# Minocycline in Acute Traumatic Spinal Cord Injury: A Systematic Review and Exploratory Meta-Analysis of Preclinical and Clinical Evidence

**DOI:** 10.64898/2025.12.19.25342695

**Authors:** Farzan Fahim, Mahsa Faramin Lashkarian, Farbod Tabasi Kakhki, Reihane Qahremani, Aydin Ghaffari, Seyed Mahrad Moosavian, Mohadese Jafari, Mahshid Ebrahimabad, Marzieh Ghasemi, Hossein Mahmoodi, Sadighe Bahmaie Kamaei, Sayeh Oveisi, Saeed Oraee Yazdani, Alireza Zali

## Abstract

**Background:** Traumatic spinal cord injury (SCI) is a major cause of long-term neurological disability, with limited pharmacological therapies targeting secondary inflammatory and neurodegenerative injury mechanisms. Minocycline, a tetracycline derivative with anti-inflammatory and neuroprotective properties, has been investigated in both experimental and clinical settings; however, its therapeutic efficacy in acute traumatic SCI remains uncertain.

**Methods:** A systematic review was conducted in accordance with PRISMA 2020 guidelines. Major electronic databases were comprehensively searched to identify preclinical animal studies and human clinical studies evaluating minocycline, alone or in combination therapies, for acute traumatic SCI. Risk of bias was assessed using Joanna Briggs Institute (JBI) critical appraisal tools tailored to study design. Qualitative synthesis included all eligible studies, while quantitative synthesis was restricted to clinical studies reporting extractable effect estimates for neurological improvement.

**Results:** A total of 11 studies met inclusion criteria for qualitative synthesis, including experimental animal studies and human clinical investigations. Preclinical studies demonstrated consistent biological effects of minocycline on inflammatory markers, oxidative stress, and histopathological outcomes, particularly in combination therapies, although functional recovery with minocycline monotherapy was inconsistent. Clinical studies indicated that minocycline was generally well tolerated; however, most trials did not demonstrate statistically significant improvements in neurological or functional outcomes. Only two clinical studies provided suitable data for meta-analysis, yielding a pooled odds ratio of 1.70 (95% CI 0.95–3.06) for neurological improvement, which did not reach statistical significance.

**Conclusion:** Current evidence suggests that while minocycline exhibits promising biological activity and an acceptable safety profile in acute traumatic SCI, robust clinical efficacy has not been conclusively demonstrated. Well-designed, adequately powered randomized controlled trials with standardized outcome reporting are required to determine whether these biological effects translate into meaningful functional recovery.

## Introduction

Traumatic spinal cord injury (SCI) is a severe neurological condition associated with substantial global health and socioeconomic burden. The worldwide incidence of traumatic SCI is estimated at 13–40 cases per million population annually, accounting for approximately 250,000–500,000 new cases each year (1, 2). Although SCI has traditionally affected younger individuals, recent epidemiological trends indicate a growing proportion of elderly patients, largely due to low-energy falls (3). Traumatic SCI disrupts supraspinal control of spinal circuitry, resulting in profound motor and sensory impairments. Clinically, affected individuals may develop spastic paralysis, complete or partial sensory loss, and dysesthetic sensations below the level of injury. Beyond the acute neurological deficits, patients frequently experience chronic secondary complications, including spasticity, neuropathic pain, autonomic dysfunction, urinary and bowel incontinence, pressure ulcers, and osteoporosis(2). The economic burden of SCI is considerable, with estimated lifetime costs per individual ranging from 0.7 to 2.5 million U.S. dollars, particularly among patients injured at a younger age, those with higher neurological levels of injury, and those requiring intensive long-term care (4, 5). Despite advances in acute care, traumatic SCI remains a major clinical challenge because no pharmacological therapy has demonstrated clear and sustained neuroprotective efficacy capable of limiting secondary injury or promoting meaningful neurological recovery.

Current management of acute traumatic SCI focuses primarily on minimizing secondary injury through early surgical decompression and spinal stabilization, which are recommended to reduce ongoing mechanical damage to the spinal cord (6-9). While these interventions address the primary insult, they do not reverse established neurological deficits or adequately prevent the cascade of secondary injury mechanisms, including inflammation, oxidative stress, and apoptosis (2, 10, 11). Methylprednisolone sodium succinate (MPSS), once widely used as an adjunctive pharmacological treatment, has demonstrated only modest and inconsistent benefits in motor function preservation and is associated with significant adverse effects, such as increased infection risk and gastrointestinal complications (12). Subsequent studies have failed to confirm its clinical efficacy, leading to a global decline in its use (13). Given the absence of consistent long-term functional or quality-of-life benefits, major clinical guidelines now advise against routine MPSS administration in acute SCI management (14). Consequently, there has been increasing interest in alternative pharmacological strategies that target specific secondary injury pathways, particularly neuroinflammation and microglial activation.

Minocycline, a second-generation tetracycline antibiotic, has emerged as a candidate neuroprotective agent due to its pleiotropic biological effects independent of its antimicrobial properties. Experimental studies have demonstrated that minocycline can inhibit microglial activation, suppress the production of pro-inflammatory cytokines such as tumor necrosis factor-α (TNF-α) and interleukin-6 (IL-6), reduce oxidative stress, and attenuate apoptotic signaling pathways following central nervous system injury (15–20). Given the critical role of microglia and inflammatory mediators in shaping tissue injury and repair after SCI, modulation of these pathways by minocycline provides a biologically plausible rationale for its therapeutic investigation in this context (19,20). Moreover, advances in drug delivery approaches, including nanoparticle-based co-delivery systems, have further expanded interest in minocycline, particularly in combination with established anti-inflammatory agents such as methylprednisolone (28,29).

However, despite compelling mechanistic justification, the evidence for minocycline efficacy in SCI remains inconsistent across preclinical and clinical studies. Several experimental studies have failed to demonstrate meaningful neuroprotection or functional recovery with minocycline monotherapy in animal models of cervical and thoracic SCI (31), whereas others have reported favorable biochemical or histological effects, particularly when minocycline is administered in combination with agents such as methylprednisolone or FK506 (28–30,32). These findings raise important questions regarding the context-dependency of minocycline’s therapeutic effects, including dosing, timing of administration, injury severity, and the potential necessity of combination therapy to achieve clinically relevant benefit.

In the clinical setting, evidence remains limited and inconclusive. A phase II randomized controlled trial investigating intravenous minocycline in acute traumatic SCI demonstrated biological activity but did not establish definitive functional superiority over placebo (21). Subsequent analyses focusing on cerebrospinal fluid and serum biomarkers suggested potential modulation of inflammatory and neuroaxonal injury markers, yet the clinical significance of these findings remains uncertain (26,27). More recent pilot studies have explored combined pharmacological approaches, including minocycline with methylprednisolone, reporting signals of neurological improvement; however, these studies are constrained by small sample sizes, heterogeneous dosing regimens, and variability in outcome assessment (22). Collectively, these limitations hinder clear interpretation of minocycline’s clinical efficacy and translational relevance.

Given the persistent uncertainty surrounding the role of minocycline in SCI, a comprehensive synthesis of both preclinical and clinical evidence is warranted. The present systematic review and meta-analysis aimed to critically evaluate the efficacy and safety of minocycline in experimental animal models and human patients with acute SCI. By integrating and stratifying evidence across study designs, this review seeks to clarify the extent to which preclinical findings translate into clinical outcomes, identify sources of heterogeneity, and inform future research directions regarding the potential role of minocycline, alone or in combination with other agents, in the management of acute SCI.

## Methodology

This systematic review and meta-analysis was conducted in accordance with the *Preferred Reporting Items for Systematic Reviews and Meta-Analyses (PRISMA) 2020* guidelines (23). To ensure methodological transparency and avoid unnecessary duplication, the protocol was prospectively registered in the International Prospective Register of Systematic Reviews (PROSPERO) on 18 November 2025. The full protocol is publicly available in PROSPERO (Registration ID: CRD420251185674).

See Supplementary File 1,2 for PRISMA checklist and study protocol.

### Search Strategy

A comprehensive literature search was conducted in November 2025 across the following electronic databases: PubMed, Scopus, Web of Science, Embase, and the Cochrane Library. No restrictions were applied regarding language or publication date to ensure broad coverage of relevant literature. Articles published in languages other than English were to be translated if included, although no such articles were ultimately retrieved.

The search included the following keywords: Minocycline; Spinal Cord Injury; Traumatic Spinal Cord Injury; Neuroprotective Therapy; Corticosteroids; Methylprednisolone.

An example of the PubMed search query is presented below:

(“Minocycline”[MeSH Terms] OR Minocin[tiab] OR “Minocin MR”[tiab] OR “Minocycline Hydrochloride”[tiab] OR “Hydrochloride, Minocycline”[tiab] OR Minocycline[tiab] OR Minakne[tiab] OR Arestin[tiab] OR Dynacin[tiab] OR Lederderm[tiab] OR Blemix[tiab] OR Cyclomin[tiab] OR Cyclops[tiab] OR Dentomycin[tiab] OR Klinomycin[tiab] OR Minoclir[tiab] OR Minolis[tiab] OR Minoplus[tiab] OR Mestacine[tiab] OR Minomycin[tiab] OR Minotab[tiab] OR Mynocine[tiab] OR “Minocycline derivative”[tiab] OR “Minocycline therapy”[tiab] OR “Minocycline treatment”[tiab] OR “Minocycline administration”[tiab]) AND (“Spinal Cord Injuries”[MeSH Terms] OR “Spinal Cord Injury”[tiab] OR “Spinal Cord Injuries”[tiab] OR “Spinal Injury”[tiab] OR “Spinal Injuries”[tiab] OR “Spinal Cord Trauma”[tiab] OR “Traumatic Myelopathy”[tiab] OR “Myelopathy, Traumatic”[tiab] OR “Post-Traumatic Myelopathy”[tiab] OR “Spinal Cord Contusion”[tiab] OR “Spinal Cord Laceration”[tiab] OR “Spinal Cord Transection”[tiab] OR “Spinal Cord Damage”[tiab] OR “Spinal Cord Lesion”[tiab] OR “Spinal Cord Compression”[tiab] OR “Traumatic Spinal Cord Injury”[tiab] OR “Acute Spinal Cord Injury”[tiab] OR “SCI”[tiab] OR (“Spinal Cord”[tiab] AND Injury[tiab]))

The complete search strategies for all databases are provided in Supplementary File 3.

Additionally, the reference lists of all eligible studies and relevant review articles were manually screened, which resulted in the identification of one additional study that was not retrieved through the initial database search but met the inclusion criteria and was therefore included in the final analysis.

### Eligibility Criteria

#### Study Design and Limits

We initially aimed to include human studies only. However, because combination therapy with minocycline (e.g., minocycline plus methylprednisolone or other agents) is an emerging field with limited clinical evidence, high-quality animal studies were also included to capture the full scope of available data. Notably, previous systematic reviews have already evaluated minocycline monotherapy in animal models; therefore, this review focused specifically on studies assessing combination therapy.

The following limits and filters were applied:

**Language:** no language restrictions.

**Population type:** both human and animal studies were eligible.

**Date range:** no date restrictions (from database inception to the date of search).

#### PICO Framework

##### Population (P)

Human participants or animal models with acute traumatic spinal cord injury. Studies involving non-traumatic etiologies (e.g., demyelinating diseases, tumors, infections) or chronic SCI were excluded.

##### Intervention (I)

Minocycline therapy administered in the acute phase of injury, either as monotherapy, or in combination with another agent, such as methylprednisolone (corticosteroid) or other adjunctive treatments.

##### Comparator (C)

Any control group not receiving minocycline, including placebo, standard care, or alternative therapies.

##### Outcomes (O)

Studies assessing at least one of the following were eligible:

Neurological outcomes, including motor or sensory function recovery

Functional improvement

Hospitalization-related outcomes, such as duration of stay (for human studies)

Any validated neurobehavioral or functional neurological score (for animal studies).

#### Exclusion Criteria

Studies were excluded if they met any of the following:

1. **Study design–related exclusions:** Case reports or case series Reviews, meta-analyses, conference abstracts, letters, or book chapters Studies without available full text
2. **Population or condition–related exclusions:** Studies conducted exclusively in chronic spinal cord injury models Studies involving non-traumatic causes of spinal cord injury (e.g., demyelinating diseases, ischemic injury) Studies investigating traumatic brain injury (TBI) instead of spinal cord injury
3. **Intervention–related exclusions:** Studies assessing corticosteroid or methylprednisolone monotherapy without any minocycline arm
4. **Outcome–related exclusions:** Studies not reporting outcomes relevant to this review, including neurological recovery, sensory or motor improvement, hospitalization duration, or functional neurologic decline (FND).

#### Screening and Selection

A total of 1,530 records were identified through database searches (PubMed: 210; Scopus: 572; Web of Science: 429; Embase: 306; Cochrane Library: 13). All records were imported into Rayyan software for duplicate removal, resulting in 823 unique publications for screening.

Title and abstract screening were independently conducted by AG, MM, and MF. Discrepancies were resolved through discussion, and unresolved conflicts were adjudicated by a fourth reviewer (FT). An Excel-based exclusion sheet, developed by FF, was used to document excluded studies, including: article title, first author, country of origin, year of publication, DOI, and reason for exclusion (see Supplementary File 4).

Following initial screening, 26 studies were retained for full-text assessment. Manual screening of reference lists identified one additional eligible study, which was included in the final analysis.

Full-text screening was independently performed by ME and RQ. Disagreements were resolved by discussion or consultation with a third reviewer (FT). Ultimately, 11 studies met all inclusion criteria and were selected for data extraction. An Excel-based inclusion and exclusion sheets, developed by FF, was used to document included and excluded studies, 2 independent included sheets contain: article title, first author, country of origin, year of publication, DOI, population, intervention, comparison group, outcome and study design; also 2 independent excluded sheets which contain: article title, first author, country of origin, year of publication, DOI and reason for exclusion are used and attached to the supplementory files. (see Supplementary File 5).

#### Data Extraction

A standardized data extraction form was developed by FF. Data extraction was independently performed by MH and AS. 2 independent extraction excel sheets are attached to supplementory file 6.

The following variables were extracted:

**Population:** Study characteristics (first author, publication year, DOI, funding source, study region, study design, center type, study period, ethical approval number, conflict of interest statement, blinding method, risk of bias), Type, Inclusion criteria, Exclusion criteria, Follow-up duration, Sample size, Age, Sex, Mechanism of injury, Completeness of injury, Neurological level of injury, Time from injury to enrollment, Baseline ASIA Impairment Scale (AIS), Baseline ASIA Motor Score, Baseline ASIA Sensory Score (Pin-prick), Baseline ASIA Sensory Score (Light touch), Baseline Spinal Cord Independence Measure (SCIM), Baseline Functional Independence Measure (FIM), Baseline Modified Rankin Scale (mRS), Other reported functional or neurological baseline measures

**Intervention:** Intervention alone or combined, Timing of initiation post-injury, Initiation setting (ER, ICU, intraoperative), Method of induction, Initial dose, Maintenance dose & duration, Time to reach target dose, Use of sedation or neuromuscular blockade, Concurrent treatments, Type of surgery, Steroid type/dosage/duration

**Comparison:** Control group treatment description

**Outcomes:** Improvement in AIS grade, change in ASIA Motor Score, Mortality rate, change in ASIA Sensory Score, Change in SCIM score, Change in FIM score, Change in WISCI II score, Change in Quality-of-Life score, Length of ICU stay, Length of hospital stay, Safety and Adverse Outcomes, Timing of Outcome Assessments, Concomitant injury, Statistical adjustment, Adverse events, Main findings, notes.

A detailed summary of all extracted is provided in Supplementary File 6.

After completing data extraction, two articles were excluded from the analysis due to statistical insufficiency, including incomplete reporting of outcome measures and lack of sufficient data for effect size calculation, which prevented their inclusion in quantitative synthesis.

#### Risk of Bias Assessment Details

Methodological quality and risk of bias were independently assessed by MM and MJ using Joanna Briggs Institute Critical Appraisal Checklists (appropriate for study design: RCT, cohort; quasi-checklist for animal studies), RCT assessment included randomization, allocation concealment, intervention delivery, blinding, outcome measurement, retention, statistical analysis, trial design; Cohort assessment included participant selection, exposure and outcome measurement, confounding control, follow-up adequacy, analysis appropriateness; Checklist items rated as Yes, No, Unclear, Not applicable. Reviewers were blinded to authorship and outcomes; Discrepancies between MM and MJ were resolved through discussion and consensus. Assessment completed over two days; 2 independent Checklists provided in Supplementary File 7.

#### Data synthesis and statical analysis

Only studies that reported extractable quantitative outcomes on AIS/Frankel grade improvement following intravenous minocycline administration for acute traumatic spinal cord injury were eligible for quantitative synthesis. After screening all available clinical and preclinical data by FF, only two human clinical studies (Casha 2012 and Meshkini 2024) provided suitable numerical effect sizes and were therefore included in the meta-analysis. Both studies directly reported odds ratios (ORs) and 95% confidence intervals (CIs) comparing neurological improvement between minocycline⍰treated patients and control groups.

Log-transformed odds ratios (log ORs) and standard errors were extracted from the two studies and pooled using a random-effects model based on restricted maximum likelihood (REML). Because only two studies were available (k=2), classical tests for publication bias, including Egger’s regression test, Begg’s rank correlation, and trim⍰and⍰fill, were not performed, as these procedures are statistically invalid for meta-analyses with fewer than three studies. A funnel plot was generated solely for descriptive visualization, without inferential interpretation.

All analyses were performed using R version 4.5.1 and the metafor package.

## Results

### 1) Study selection

The study selection process is outlined in the PRISMA 2020 flow diagram (Figure 1). The initial search identified 1,530 records across all targeted databases. After removing duplicates, 539 unique studies remained and were screened by title and abstract. Following this stage, 26 studies were retained for full-text assessment based on predefined inclusion criteria.

**Figure 1:**
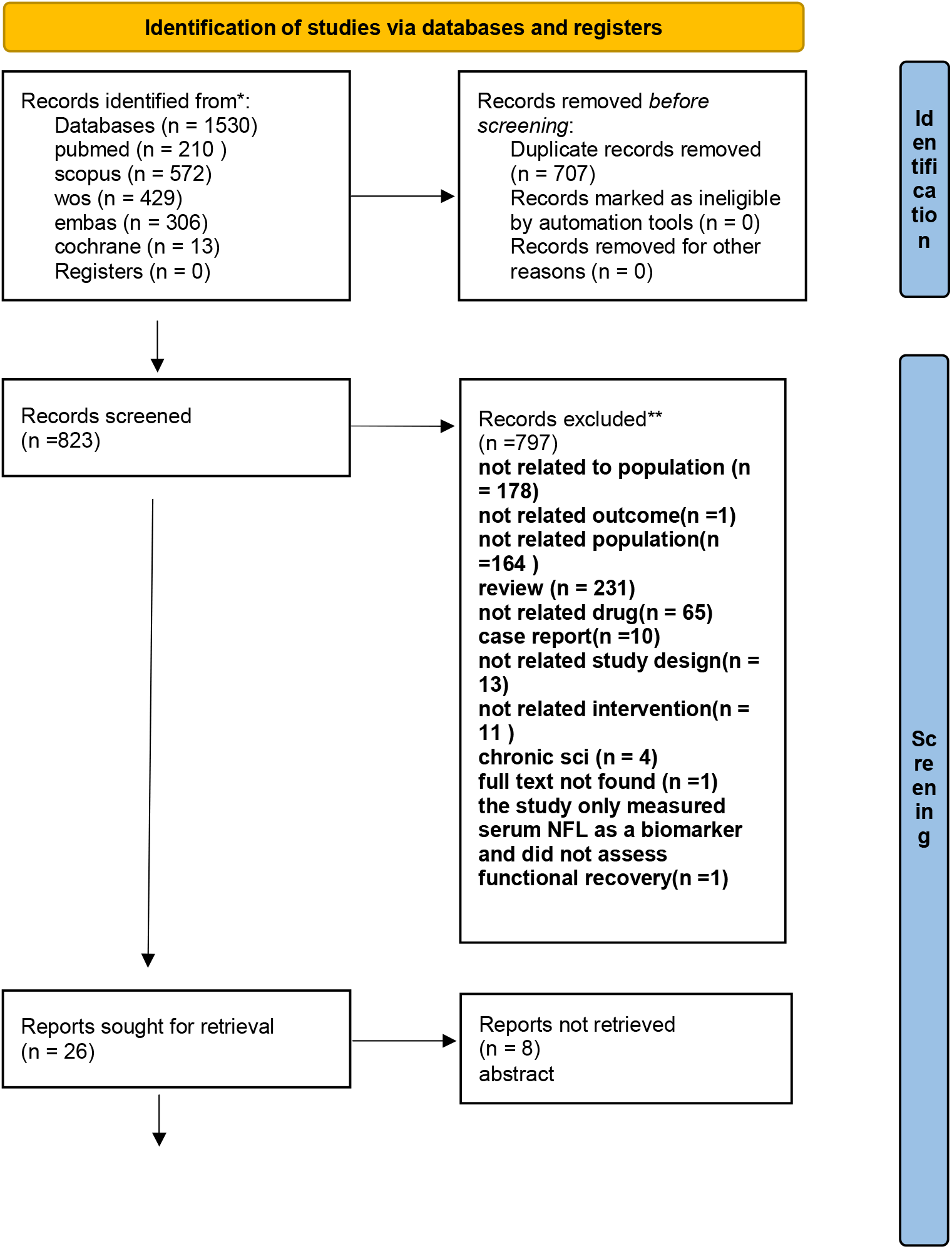
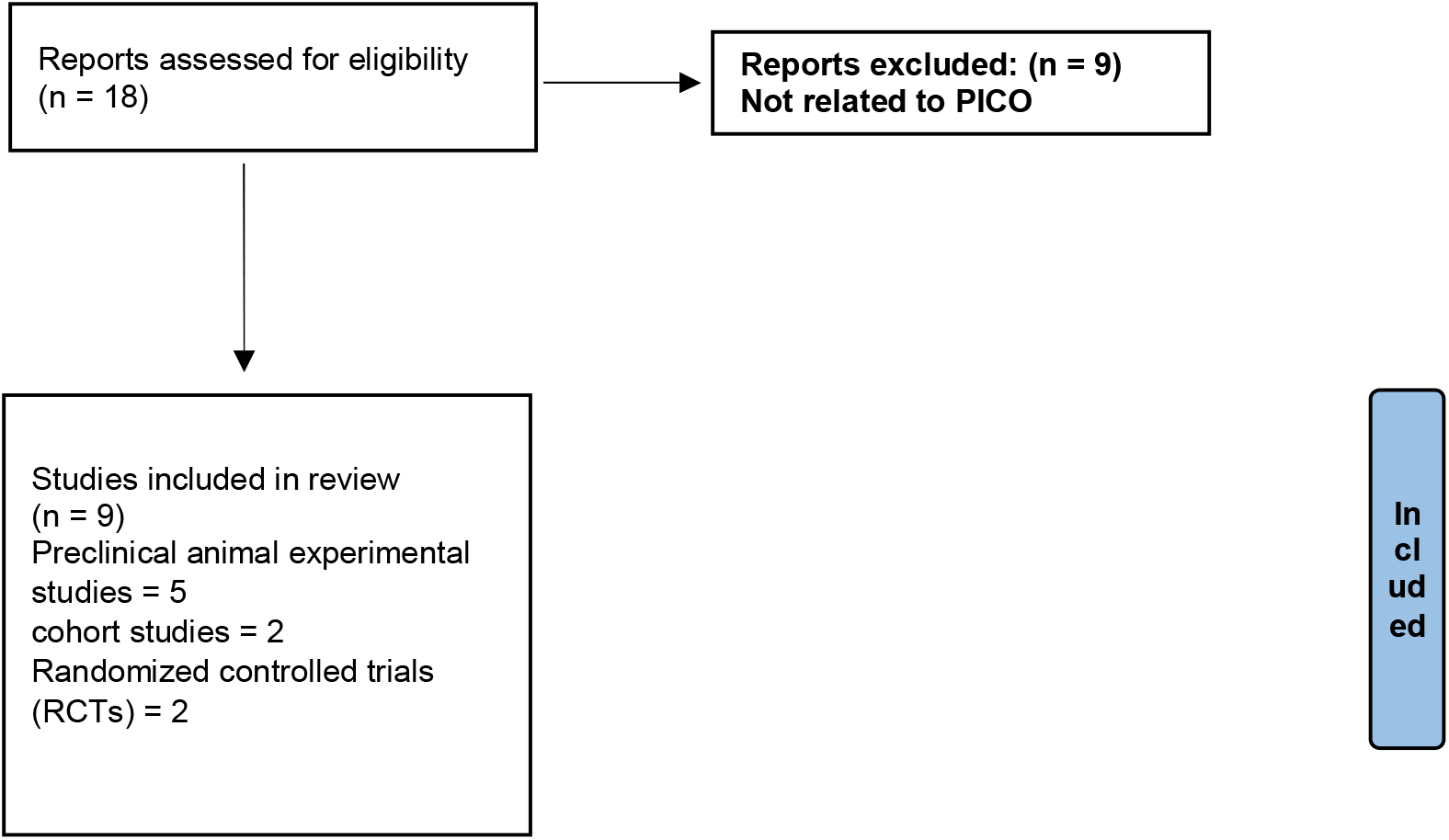
PRISMA flow diagram.

Upon full-text review, 9 studies met the eligibility criteria and were included in the final synthesis. Reasons for exclusion at this stage were documented separately. The final included studies consisted of 5 preclinical animal experimental studies,1 observational study based on previously conducted randomized clinical trial data, 3 randomized controlled trials (RCTs). All included studies investigated the therapeutic effects of minocycline in traumatic spinal cord injury (SCI).(Figure 1)

### 2) Characteristics of included studies

A total of nine eligible studies, published between 2010 and 2024, were included in the final synthesis. These studies originated from Canada (n=4), China (n=2), Turkey (n=1), Saudi Arabia (n=1), and Iran (n=1). Five studies were preclinical experimental investigations in animal models, two were randomized controlled trials (RCTs), and two were clinical cohort or observational analyses derived from randomized trial datasets. Sample sizes ranged from 32 to 94 participants in clinical studies and 30 to 80 animals in preclinical experiments. Substantial heterogeneity was observed across studies with respect to minocycline dosage, route of administration, and treatment duration. Interventions included intraperitoneal administration in animal models, intravenous administration in human trials, and oral or nanoparticle-based delivery approaches in experimental settings. Follow-up durations varied considerably, ranging from 72 hours in biomarker-focused clinical studies to 24 months in long-term assessments of neurological and functional recovery. (Table 1)

**Table 1.**
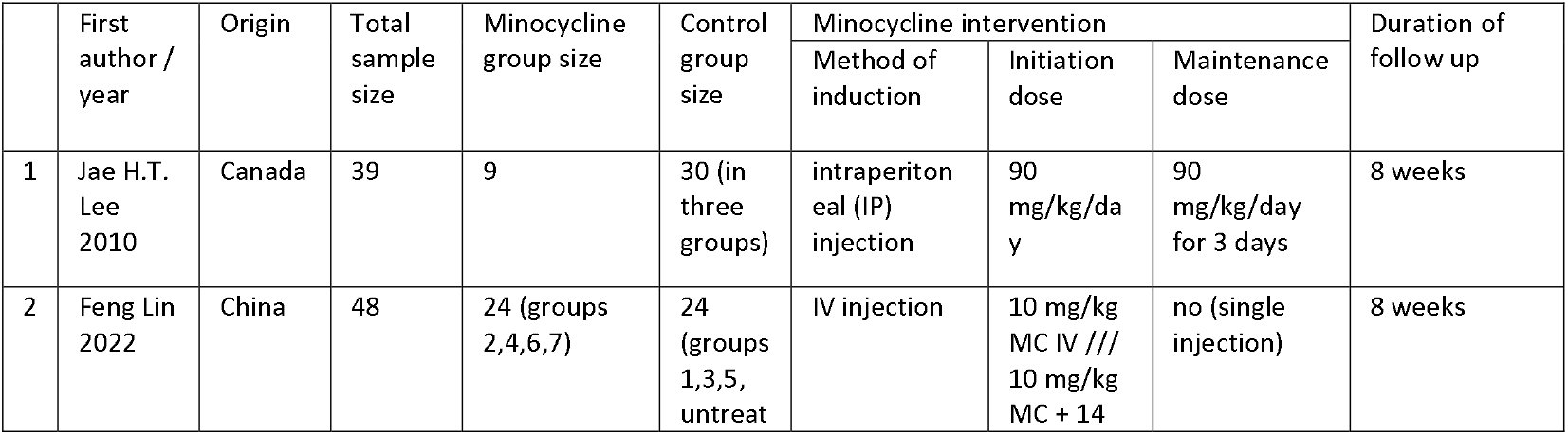
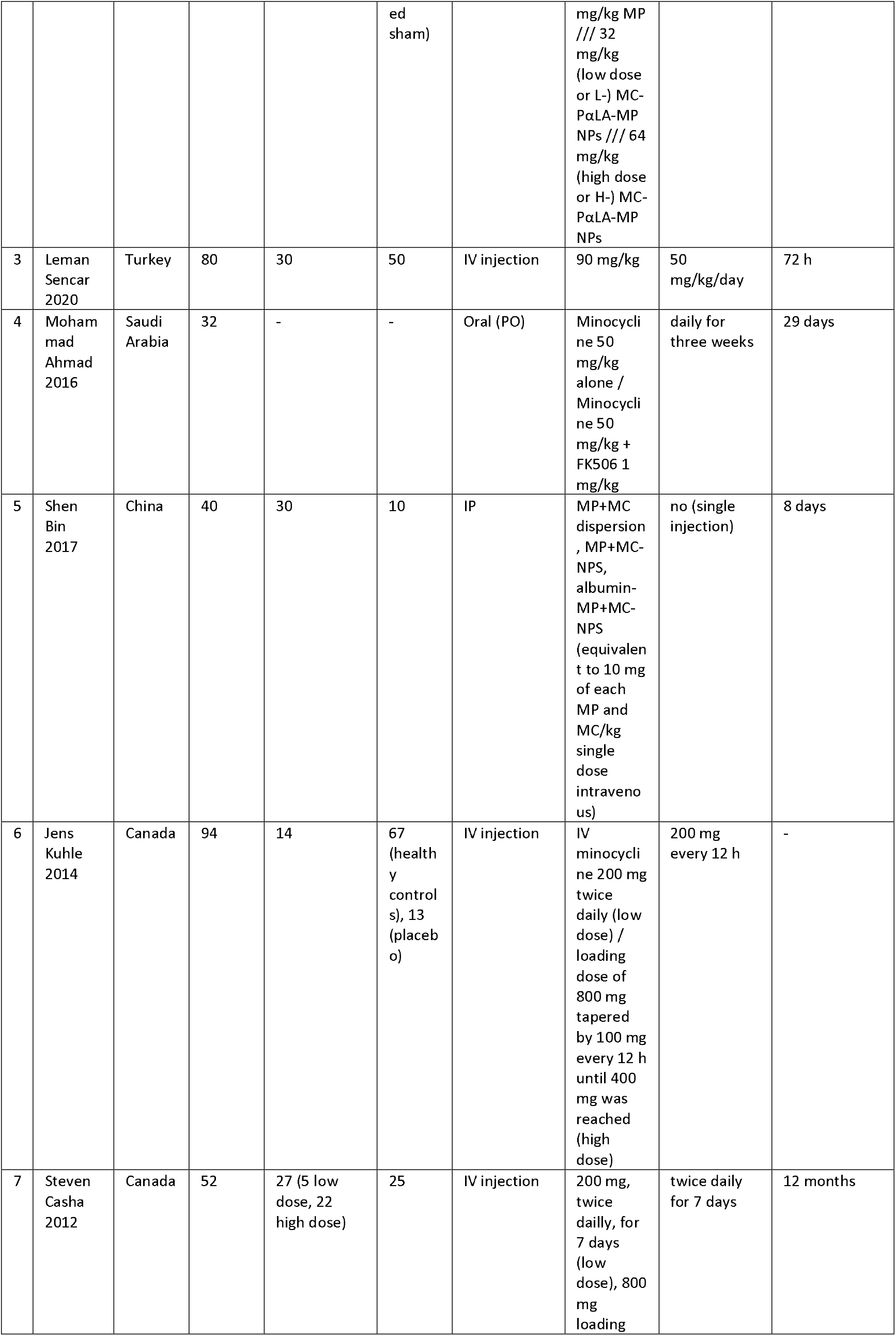
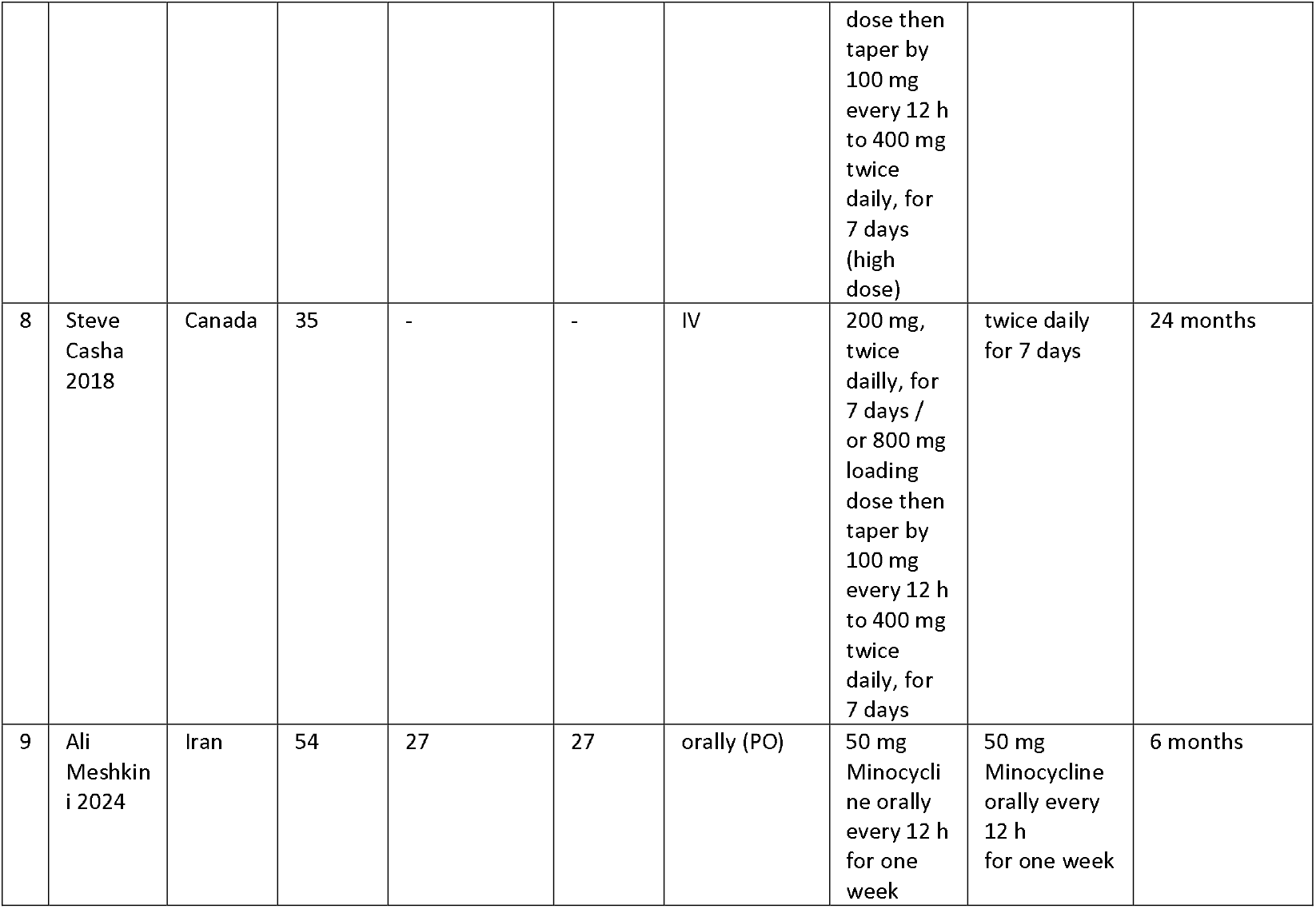
Characteristics of included studies.

### 3) Risk of Bias Assessment

Overall risk of bias for the included studies was assessed using the Joanna Briggs Institute (JBI) Critical Appraisal Tools, with checklist selection tailored to each study design. A summary of study-level risk of bias judgments is presented in Table□2, and domain-specific assessments are illustrated in Figures□2A–C. Among experimental animal studies, overall methodological quality was generally favorable. Three studies, Lee et□al. (2010), Ahmad et□al. (2016), and Sencar et□al. (2020), were judged to be at low risk of bias, reflecting appropriate experimental design, standardized injury induction, clearly defined intervention protocols, and comprehensive outcome assessment. Two nanoparticle-based combination therapy studies were rated as having moderate and moderate–high risk of bias, primarily due to insufficient reporting of allocation procedures, limited blinding of outcome assessment, and potential performance bias related to complex intervention delivery.Among clinical studies, one randomized controlled trial (Meshkini et□al., 2024) was judged to be at low risk of bias, supported by adequate randomization procedures, comparable baseline characteristics between groups, and complete outcome reporting. The phase□II trial by Casha et□al. (2012) was assessed as having moderate risk of bias, mainly due to limitations in blinding and sample size constraints inherent to early-phase clinical trials.The two prospective cohort studies derived from the phase□II trial dataset (Kuhle et□al., 2014) were rated as having moderate risk of bias, reflecting potential confounding, lack of adjustment for key prognostic variables, and reliance on secondary biomarker analyses rather than functional endpoints.No study was excluded from the qualitative synthesis on the basis of risk of bias alone; however, methodological limitations were carefully considered when interpreting the strength and translational relevance of the available evidence.

**Table 2.**
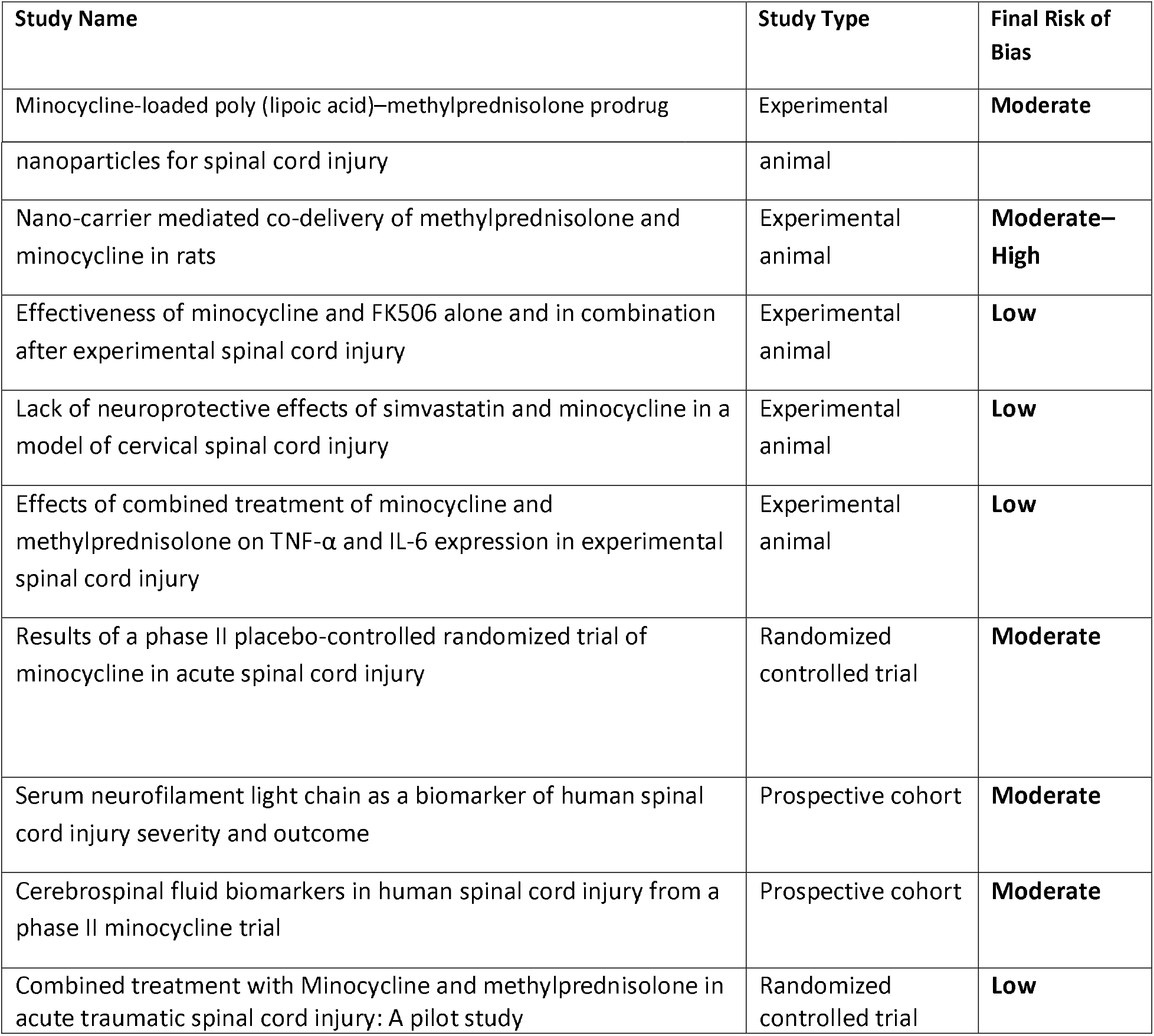
Final Risk of Bias Summary Table.

**Figure 2A:**
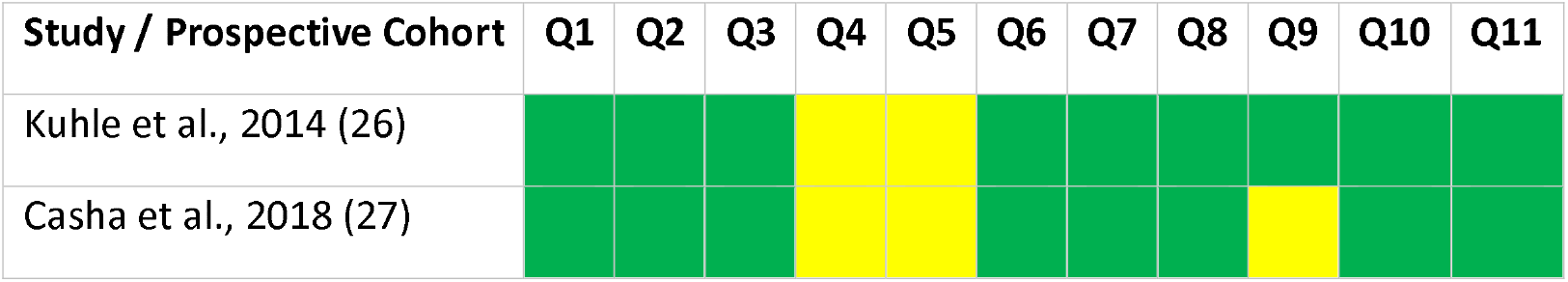
JBI Checklist for Cohort Studies (Q1–Q11)

**Figure 2B:**
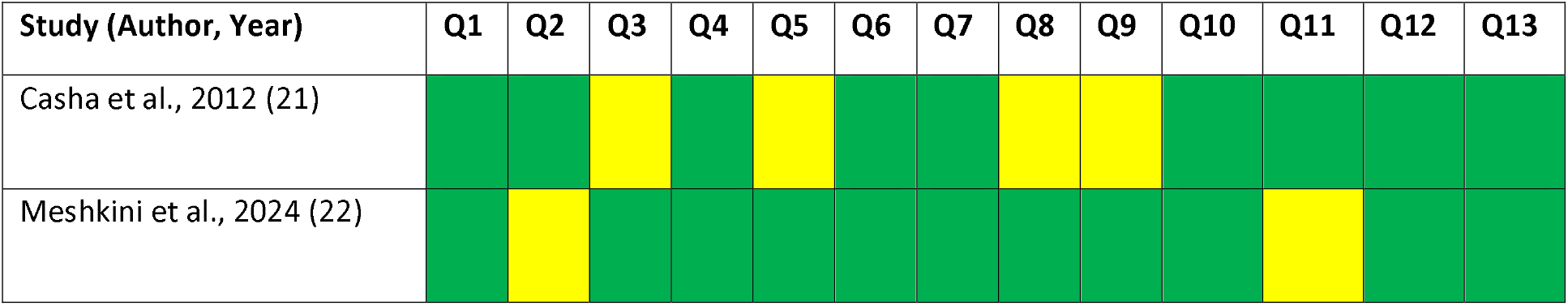
JBI Checklist for Randomized Controlled Trials (Q1–Q13)

**Figure 2C:**
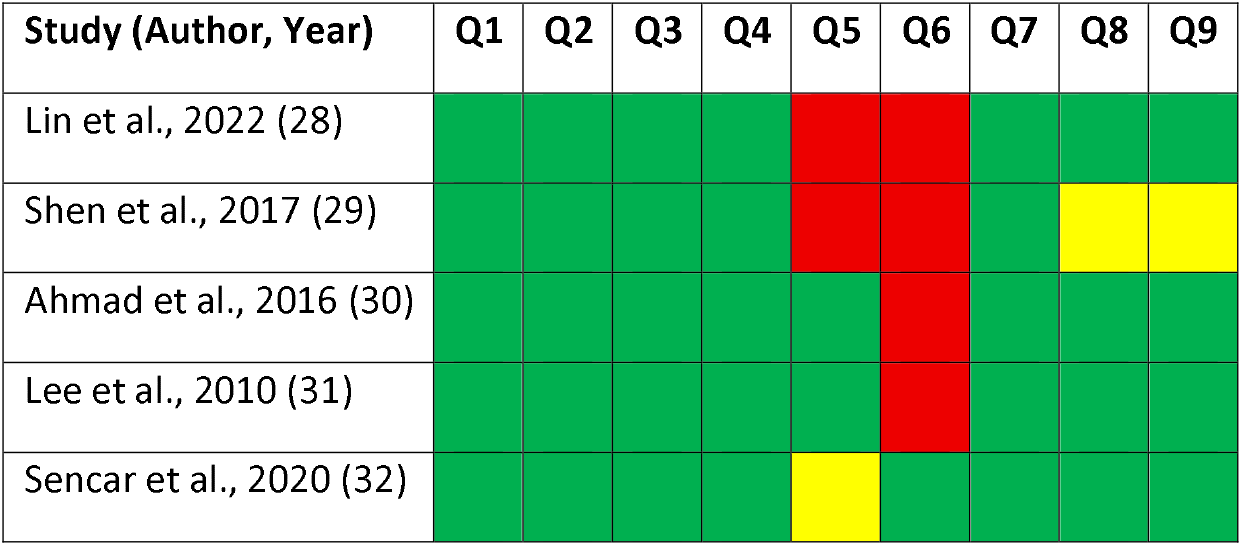
JBI Checklist for Experimental Studies (Q1–Q9)

### 4) Results of outcomes

Animal studies:

1. **Jae H. T. Lee** (31) and colleagues conducted an experimental study to evaluate the effects of adjunctive minocycline administration in a rat model of cervical spinal cord injury. A total of 39 rats were included, of which 9 received minocycline treatment and 30 served as untreated controls. Animals were followed for 8 weeks post-injury.

Functional recovery was assessed using a comprehensive behavioral test battery, including the horizontal ladder test, cylinder rearing test, modified Montoya’s staircase test, grooming test, and sensory evaluation for mechanical allodynia. For each test, outcomes were analyzed both relative to baseline performance (pre-injury) and between treatment groups.

Both groups demonstrated significant motor and sensory deficits following spinal cord injury compared with their own baseline values, confirming the success of injury induction. However, no statistically significant differences were detected between the minocycline-treated group and control group across any behavioral outcome measures during follow-up, indicating that minocycline did not confer measurable functional improvement in this study.

Additionally, histological analyses were performed to assess tissue preservation and axonal response, including spared white and gray matter quantification, as well as corticospinal (CST) and reticulospinal tract (RST) sprouting. Consistent with the behavioral outcomes, no significant differences were observed between groups in any of the histological parameters.

2. The study by **Feng Lin** et al.(28) was conducted on 48 rats subjected to spinal cord injury (SCI), followed by an 8-week evaluation period. A total of 24 rats received minocycline during the study, while 24 animals served as untreated controls. Functional recovery was assessed using the Basso, Beattie, Bresnahan (BBB) locomotor rating scale, along with measurement of bladder function recovery time. In addition, histological analysis and immunohistochemistry were performed to evaluate tissue preservation. The expression levels of pro-inflammatory cytokines, including tumor necrosis factor-α (TNF-α), interleukin-1β (IL-1β), and interleukin-6 (IL-6), were quantified in the supernatants of injured spinal cord tissue lysates.

This study was primarily designed to examine the efficacy of nanoparticle-based drug delivery to the central nervous system. Therefore, eight experimental groups were included for comparison. To assess the therapeutic effect of minocycline independently (without nanocarriers), three drug-only groups were evaluated: minocycline monotherapy, methylprednisolone monotherapy, and a combination of both agents.

Across these three groups, no statistically significant differences in locomotor recovery or pro-inflammatory factors were reported. Although the article did not provide exact numerical values for all comparisons, examination of the presented charts indicated no significant differences in outcomes among the groups.

3. **The study by Leman Sencar et⍰al. (32)**

The study by Leman Sencar and colleagues was designed to evaluate the effects of combined minocycline and methylprednisolone (MP) treatment on inflammatory responses following spinal cord injury (SCI), with a particular focus on TNF-α and IL-6 expression. The combined regimen was compared with each agent administered alone. A total of 80 rats were randomized into six experimental groups, of which three key groups (n⍰=⍰15 per group) received minocycline alone, MP alone, or minocycline + MP and were included in this review (Table⍰3).

**Table 3.** Effect on Tissue Injury, Edema & Cavitation.

|  | Effect on Tissue Injury, Edema & Cavitation |
| --- | --- |
| MC | Significant lower protection compared to two other groups at 24 h, more protective effect at 72 h |
| MP | Significant structural protection at 24 h, preserved at 72 h |
| MP+MC | Best preservation and significant reduced edema at both timepoints |

Across all evaluated outcome domains, the combined minocycline + MP regimen consistently demonstrated superior effects compared with either monotherapy. This included greater reductions in inflammatory markers, improved structural preservation, and enhanced functional recovery measures. While both monotherapy groups showed partial biological activity, neither achieved improvements comparable to those observed with the combined treatment.

- **Inflammatory Outcomes (TNF-α and IL-6 Expression):** At 24⍰hours post-injury, animals treated with minocycline (MC) and methylprednisolone (MP) showed a significant increase in pro-inflammatory cytokine expression. The combined treatment group also demonstrated an early increase in TNF-α and IL-6 expression; however, this increase was not statistically significant compared with the control group.By 72⍰hours post-injury, all three treatment groups exhibited reduced TNF-α and IL-6 expression relative to the 24-hour time point. Nevertheless, cytokine levels remained significantly higher than those observed in the control group. Among the treatment strategies, the combination therapy produced the greatest and most sustained reduction in cytokine expression at both time points.
- **Histopathological Outcomes:** Evaluation of tissue injury demonstrated time-dependent differences among treatment groups. Minocycline treatment was associated with delayed neuroprotection, characterized by reduced edema, hemorrhage, and cavitation primarily at later time points. MP treatment resulted in more pronounced early tissue preservation, whereas the combination therapy provided the most extensive overall protection, with improved neural tissue integrity observed at both 24 and 72⍰hours. Because tissue damage severity was assessed using observer-based methods, statistical comparisons between groups were not performed.
- **Oxidative Stress Markers:** Both monotherapy groups demonstrated significantly increased levels of malondialdehyde (MDA) and decreased levels of superoxide dismutase (SOD) compared with the control group. In contrast, the combination therapy group exhibited smaller changes in MDA and SOD levels than either monotherapy group, although these values remained significantly different from those of the control group.

**4. The study by Mohammad Ahmad et⍰al. (30)** investigated the therapeutic effects of minocycline (MC) on spinal cord injury (SCI) using 32 rats with a 29-day follow-up period. Although the total sample size was reported, the exact number of animals receiving MC monotherapy was not clearly specified. The study compared minocycline monotherapy, FK506 (tacrolimus) monotherapy, combined minocycline + FK506 therapy, and untreated SCI controls. A comprehensive range of outcomes was assessed, including behavioral measures (BBB locomotor score, Tarlov score, inclined-plane test, and functional deficit scoring), histopathological analysis, and biochemical parameters (spinal monoamines [5-HT and 5-HIAA], lipid peroxidation levels, glutathione [GSH] activity, and myeloperoxidase [MPO] activity). Overall, minocycline monotherapy consistently demonstrated greater neuroprotective effects than FK506 alone across assessed outcome domains. The combined minocycline + FK506 treatment produced the greatest overall improvements relative to either monotherapy.

- Behavioral outcomes: Across all behavioral assessments, including BBB scoring, Tarlov score, inclined-plane performance, and functional deficit scoring, minocycline treatment resulted in significant functional improvement compared with the untreated SCI group and showed consistently greater improvement than FK506 alone. The combined treatment group demonstrated superior behavioral recovery compared with both monotherapy groups.
- **Histopathological outcomes:** Minocycline treatment was associated with better preservation of spinal cord tissue compared with FK506 monotherapy. However, the combination therapy produced the most pronounced tissue preservation among all experimental groups.
- **Biochemical outcomes:** Minocycline monotherapy exerted measurable antioxidant and anti-inflammatory effects. In comparison, the combined treatment group showed greater improvements across biochemical markers, indicating a more favorable overall biochemical profile than either monotherapy.

**5. The study by Shen⍰Bin et⍰al. (29)** was conducted on 40 rats and primarily evaluated the effectiveness of nanoparticle-based delivery systems for minocycline (MC) and methylprednisolone (MP) following spinal cord injury (SCI). The study did not assess minocycline monotherapy; instead, different delivery strategies for combined MP + MC treatment were compared. The findings indicate that treatment outcomes varied substantially according to the drug-delivery method.

- **Anti-inflammatory outcomes:** Combined MP and MC administered as free drug dispersion reduced TNF-α and IL-1β levels relative to untreated SCI controls, although the magnitude of reduction was smaller than that observed with nanoparticle-based formulations: Combined MP and MC delivered via non-targeted nanoparticles (MP + MC–NPS) produced greater reductions in inflammatory cytokines compared with the free-drug formulation. The albumin-targeted MP + MC nanoparticle formulation (Albumin–MP + MC–NPS) demonstrated the largest reductions in TNF-α and IL-1β among all treatment groups. Overall, reductions in pro-inflammatory cytokines were observed across all MP + MC treatment groups, with the greatest reductions associated with albumin-targeted nanoparticle delivery.
- **Behavioral outcomes:** Animals treated with free Combined MP and MC exhibited modest functional improvements compared with untreated SCI controls. MP + MC–NPS treatment resulted in greater behavioral recovery than the free-drug formulation. The Albumin–MP + MC–NPS group showed the most pronounced functional improvements across all assessed post-injury time points.
- **Histological outcomes:** Free MP + MC treatment was associated with a moderate reduction in lesion volume. Greater pseudocyst reduction was observed in the MP + MC– NPS group compared with the free-drug formulation. The Albumin–MP + MC–NPS group demonstrated the smallest lesion volumes and the greatest preservation of spinal cord morphology.
- **Cytotoxicity and safety outcomes:** Nanoparticle-based MP + MC formulations showed low cytotoxicity at therapeutic doses. In contrast, free MP + MC treatment was associated with higher toxicity at increased doses, whereas nanoparticle delivery mitigated dose-related toxicity.

Human studies:

**6. The study by Jens⍰Kuhle et⍰al. (26)** investigated serum neurofilament light chain (NfL) concentrations as a biomarker of neuronal injury and spinal cord injury (SCI) severity. Serum samples were obtained from participants enrolled in a phase II randomized, placebo-controlled clinical trial evaluating minocycline treatment in acute SCI. When all SCI subtypes treated with minocycline were analyzed together, minocycline administration did not result in a statistically significant reduction in serum NfL levels compared with placebo. Consistent with this finding, no significant differences were observed between treatment groups in clinical outcome measures, including ASIA motor scores, pinprick sensation, and light-touch scores. Subgroup analysis of patients with motor-complete SCI (cSCI; n⍰=⍰13) identified lower serum NfL concentrations in the minocycline-treated group (n⍰=⍰6) compared with placebo-treated patients (n⍰=⍰7). However, no corresponding statistically significant differences were detected between groups in clinical outcome measures within this subgroup. Dose-stratified analyses further showed greater reductions in serum NfL levels among patients receiving high-dose minocycline compared with placebo. As observed in the primary and subgroup analyses, these biomarker changes were not accompanied by measurable improvements in clinical outcomes.

**7. The study by Steven⍰Casha et⍰al. (21)** was a single-centre, double-blind, randomized, placebo-controlled phase II clinical trial designed to evaluate the effects of minocycline compared with placebo in patients with spinal cord injury (SCI). A total of 52 patients were included, with 27 allocated to the minocycline group and 25 to the placebo group. Among patients receiving minocycline, five received a low dose and twenty-two received a high dose. Neurological recovery was assessed using American Spinal Cord Injury Association (ASIA) motor and sensory scores over a 12-month follow-up period. Patients treated with minocycline demonstrated a mean increase of six ASIA motor points compared with placebo (95% CI, 3–14); however, this difference did not reach statistical significance (P⍰=⍰0.20). In subgroup analyses of patients with cervical SCI (n⍰=⍰25), minocycline treatment was associated with a greater mean improvement in ASIA motor score compared with placebo (mean difference 14 points; 95% CI, 0–28). Within this subgroup, numerically larger motor score differences were observed in patients with motor-incomplete injuries relative to those with motor-complete injuries, although none of these subgroup comparisons reached statistical significance. Sensory outcomes showed a similar pattern. Improvements in ASIA pinprick and light-touch scores were numerically greater in the minocycline group compared with placebo; however, these differences were not statistically significant (pinprick: mean difference 9 points, P⍰=⍰0.15; light touch: mean difference 7 points, P⍰=⍰0.27). Functional recovery was assessed using the Spinal Cord Independence Measure, Functional Independence Measure, London Handicap Scale, and Short Form-36. Across all functional outcome measures, patients receiving minocycline demonstrated numerically greater improvements compared with placebo, but no statistically significant differences were detected between groups.

**8. In a study by Steve⍰Casha et⍰al. (27)**, the effects of minocycline on cerebrospinal fluid (CSF) biomarkers following acute spinal cord injury (SCI) were evaluated. The biomarkers analyzed included interleukin-1β (IL-1β), matrix metalloproteinase-9 (MMP-9), C-C motif chemokine ligand 2 (CCL2), C-X-C motif chemokine 10 (CXCL10), neurofilament heavy chain (NfH), heme oxygenase-1 (HO-1), neural cell adhesion molecule (NCAM), and nitric oxide oxidation products (NOx). Serial CSF samples were obtained from 29 patients enrolled in a phase II randomized, placebo-controlled trial investigating intravenous minocycline in acute traumatic SCI. Patients were allocated to receive either minocycline or placebo. Comparative analyses between the minocycline-treated and placebo-treated groups did not demonstrate statistically significant differences in most CSF biomarkers. Although early between-group differences were observed in NfH levels, these differences were not statistically significant and were not maintained at later time points. In contrast, HO-1 concentrations increased at day⍰7 in placebo-treated patients, whereas this delayed increase was attenuated in the minocycline group. This difference between treatment groups reached statistical significance (P⍰=⍰0.013). HO-1 was the only CSF biomarker demonstrating a statistically significant difference associated with minocycline treatment in this study. No formal statistical analysis was reported assessing the association between minocycline treatment and ASIA motor score as a clinical treatment effect outcome.

**9) Ali Meshkini** et al.(22) investigated the effects of minocycline in combination with methylprednisolone (MP), compared with MP alone, in patients with acute traumatic spinal cord injury (SCI) in a double-blind, single-center randomized clinical trial. A total of 54 patients (27 per group) were randomized and followed for 6 months. Neurological recovery was assessed using the Frankel Grading System at baseline and at 3 and 6 months after injury. At the 3-month follow-up, improvement to higher functional grades (Frankel D or E) was observed in 7.4% of patients treated with MP alone and in 22.2% of patients treated with combined minocycline and MP; however, this difference did not reach statistical significance (odds ratio [OR] 1.34; 95% confidence interval [CI] 0.997–1.813; *p* = 0.052). At the 6-month follow-up, a greater proportion of patients in the combination therapy group achieved improvement to Frankel grades D or E compared with the MP-only group (48.1% vs. 33.3%, respectively). Mixed-effects analysis demonstrated a statistically significant association between combination therapy and neurological improvement at 6 months (OR 1.45; 95% CI 1.074–1.952; *p* = 0.015).

In other words, the addition of minocycline to standard methylprednisolone therapy was associated with a significant improvement in neurological outcomes at 6 months, as measured by Frankel grade, whereas no significant difference was observed at 3 months. These findings suggest a potential delayed neurological benefit of minocycline when used as part of combination therapy in acute traumatic SCI.

### Quantitative Synthesis of Clinical Outcomes

Two clinical studies were included in the quantitative synthesis. Both evaluated intravenous minocycline in patients with acute cervical spinal cord injury and reported AIS/Frankel grade improvement as a dichotomous outcome. Casha (2012)(21) demonstrated a large but imprecise treatment effect (OR = 2.97; 95% CI: 1.00–8.81). Meshkini (2024)(22) reported a smaller but statistically significant improvement (OR = 1.45; 95% CI: 1.08–1.95). When pooled using a random⍰effects model, the combined effect favored minocycline but did not reach statistical significance (OR = 1.70; 95% CI: 0.95–3.06). A funnel plot of log OR versus standard error was produced; however, because only two studies contributed data, no assessment of asymmetry or publication bias was feasible (Figure 3 A,B). No additional clinical or preclinical outcomes were eligible for meta-analysis due to insufficient or incompatible numeric reporting.

**Figure 3A.**
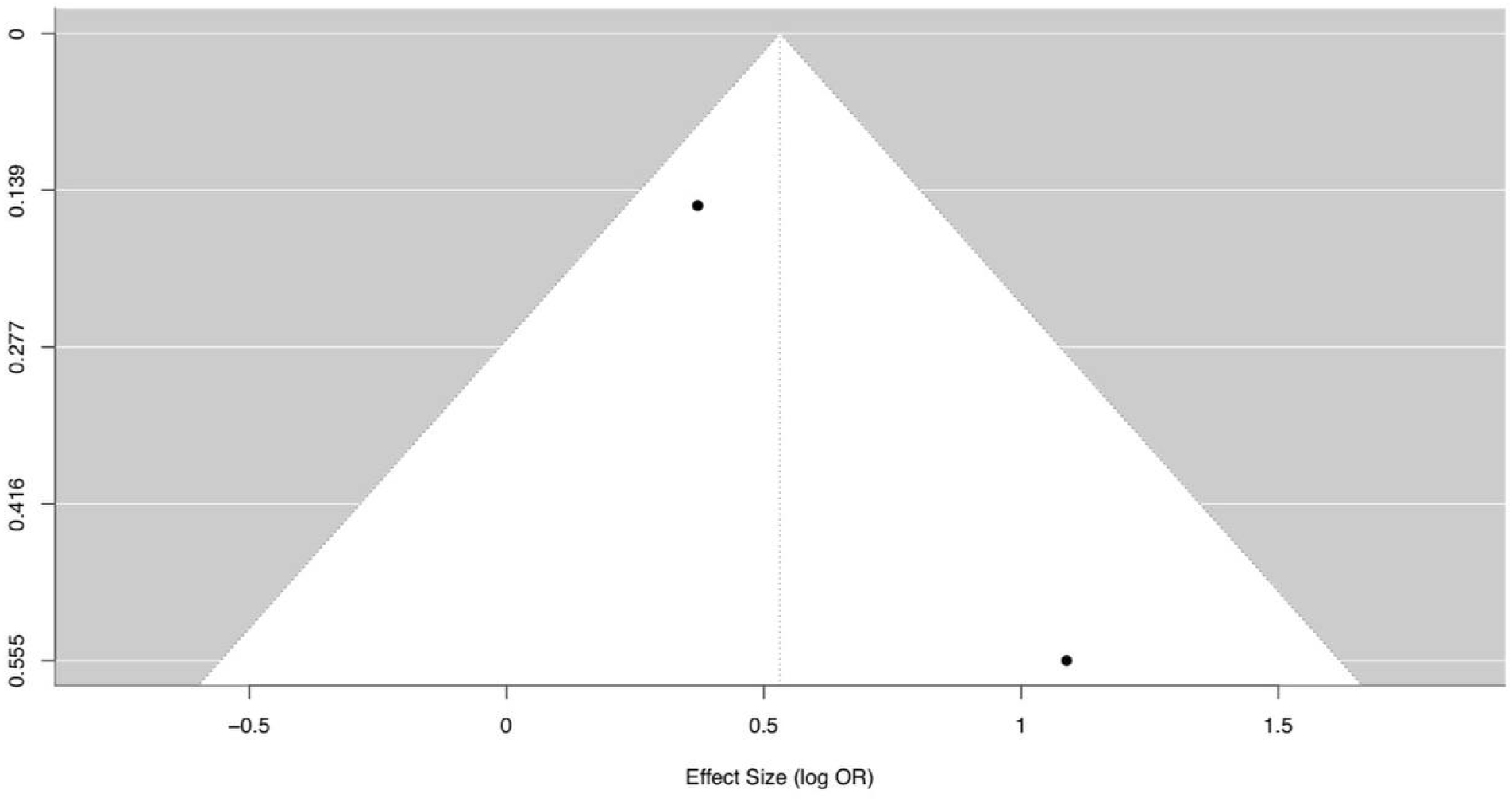
publication bias funnel plot

**Figure 3B.**
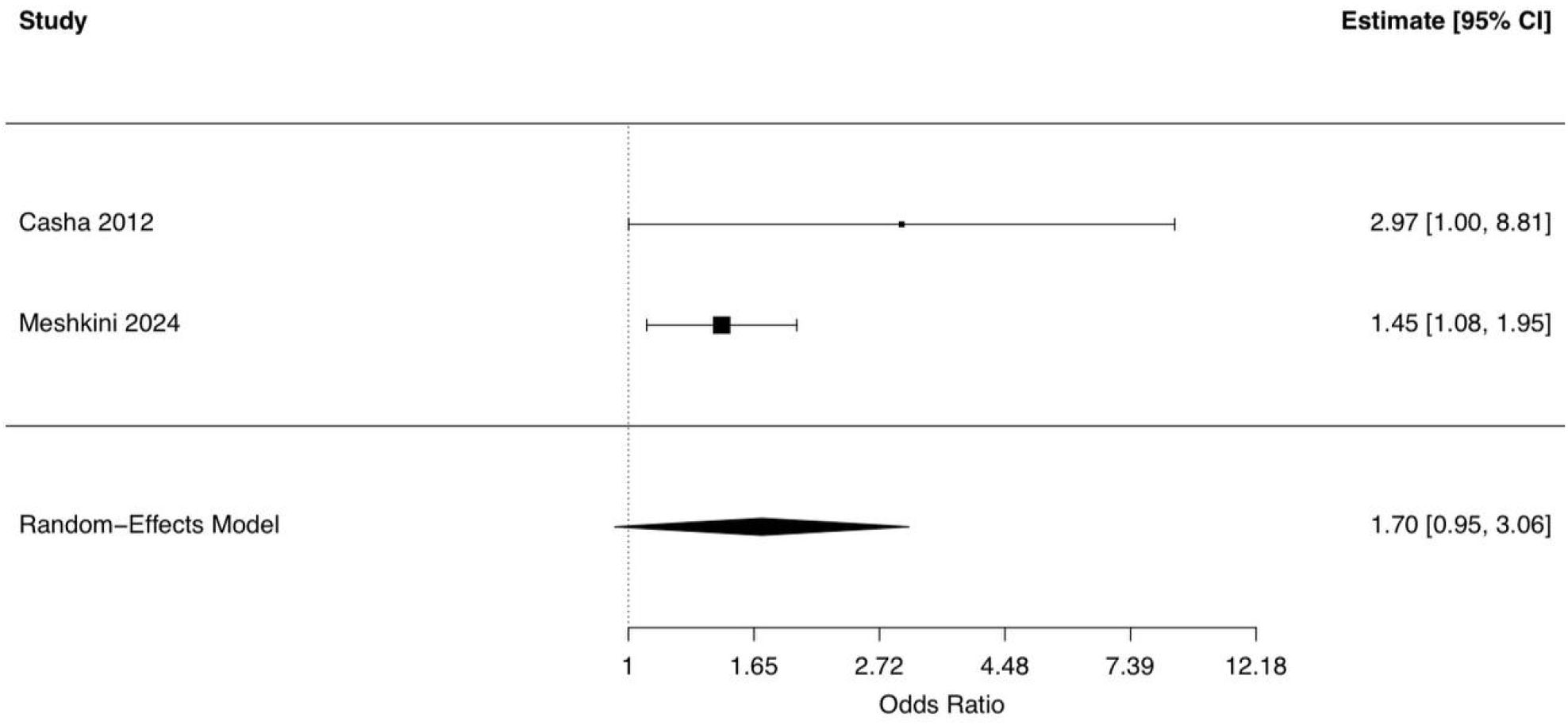
forest plot

In summary: Two clinical studies were included in the quantitative synthesis, both evaluating the effect of intravenous minocycline on AIS/Frankel grade improvement in patients with acute cervical spinal cord injury (21,22). Individual study characteristics and effect estimates are summarized in Table 4.

When pooled using a random-effects model, the combined effect favored minocycline; however, the overall result did not reach statistical significance (OR = 1.70; 95% CI: 0.95–3.06), indicating a directionally positive but inconclusive treatment effect (Figure 3B). The corresponding forest plot illustrates the variability in effect magnitude between the two contributing trials.

**Table 4.** summary of outcomes.

|  | Author | MC treated:total | Outcomes | Statistical outcome |
| --- | --- | --- | --- | --- |
| 1 | Jae H. T. Lee | 9:39 | Behavioral outcomes: | no statistically significant differences |
|  |  |  | - Horizontal ladder test<br>- Cylinder rearing test<br>- Modified Montoya's staircase test<br>- Grooming test<br>- Sensory evaluation for mechanical allodynia<br>**Histological recovery**<br>(White matter and gray matter sparing, Axonal sprouting in corticospinal and rubrospinal tracts) |  |
| 2 | Feng Lin | 24:48 | **Functional recovery**<br>(Basso, Beattie, Bresnahan (BBB) locomotor rating scale) | no statistically significant differences |
|  |  |  | **histological analysis** |  |
|  |  |  | **Immunohistochemistry** |  |
| | | | **expression levels of pro-inflammatory cytokines** (TNF-α, IL-1β, and IL-6) | |
| 3 | Leman Sencar | 30:80 | **Inflammatory Outcomes** (TNF-α & IL-6 Expression) | Significant decrease in TNF-α And IL-6 expressions in combined treatment group (MP+MC) at 24 hours after injury |
|  |  |  | **Histopathological Outcomes** | Increase in the number of inflammatory cells in the injury site<br>Decreased lipid peroxidation and neuronal and glial cell death were also observed in combined treatment group |
|  |  |  | **Oxidative Stress Markers** (MDA,SOD) | There was a significant decrease in MDA and increase in SOD levels in MP+MC group |
| 4 | Mohammad Ahmad | ? : 32 | **Behavioral outcomes:**<br>- BBB locomotor score<br>- Tarlov score<br>- Inclined plane test<br>- Functional deficits scoring | the effects of the tested drugs for SCI recovery were as follows: FK506 + minocycline > minocycline > FK506 in all studied parameters. Thus, minocycline and FK506 may prove to be a potential therapy cocktail to treat acute SCI |
|  |  |  | **histopathological outcomes** |  |
|  |  |  | **biochemical parameters:**<br>- 5-HT and 5-HIAA<br>- lipid peroxidation levels<br>- GSH activity<br>- MPO activity) |  |
| 5 | Shen Bin<br>2017 | 30:40 | **Anti-inflammatory outcomes**<br>(TNF-α, IL-1β) | Albumin-MP+MC nanoparticles were non-toxic and provided superior anti-inflammatory, neuroprotective, and functional benefits in SCI rats compared with non-targeted formulations, enabling site-specific, controlled drug delivery, reduced lesional volume, and lower effective doses |
|  |  |  | **Behavioral outcomes** |  |
|  |  |  | **Histological outcomes** |  |
|  |  |  | **Cytotoxicity and safety outcomes** |  |
| 6 | Jens Kuhle<br>2014 | 14:94 | **biomarker of neuronal damage and SCI**<br>(serum NFL concentrations) | Minocycline-treated patients exhibited significantly lower serum NFL concentrations compared with the placebo group, indicating reduced axonal injury |
| 7 | Steven Casha<br>2012 | 27:52 | **Neurological recovery**<br>(ASIA motor and sensory scores) | no statistically significant improvement |
|  |  |  | **Functional recovery**<br>- Spinal Cord Independence Measure,<br>- Functional Independence Measure<br>- London Handicap Scale<br>- Short Form-36. |  |
| 8 | Steve Casha<br>2018 | | **CSF biomarkers**<br>(IL-1β, MMP-9, CCL2, CXCL10, NfH, HO-1, NCAM, and NOx) | HO-1 was only CSF biomarker that showed a statistically significant difference |
| 9 | Ali Meshkini<br>2024 | 27:54 | **Neurological recovery**<br>(Frankel grading score) | The addition of minocycline to standard methylprednisolone therapy was associated with a significant improvement in neurological outcomes at 6 months, as measured by Frankel grade, whereas no significant difference was observed at 3 months |

Due to the inclusion of only two studies, formal assessment of publication bias was not feasible. A funnel plot was generated for descriptive purposes only and is presented in Figure 3A, without inferential interpretation. No additional clinical or preclinical outcomes were eligible for quantitative pooling because of insufficient or incompatible numeric reporting.

### Heterogeneity across Included Studies

Substantial heterogeneity was observed across the included studies with respect to study design, intervention strategy, outcome measures, and follow-up duration. Human clinical studies differed in their use of minocycline as monotherapy versus combination therapy with methylprednisolone, as well as in dosing regimens, routes of administration, and timing of treatment initiation. Outcome assessment varied considerably, with neurological recovery evaluated using ASIA motor and sensory scores, Frankel grading, or composite functional scales, limiting direct quantitative comparison across trials. Follow-up periods ranged from short-term biomarker assessment within the first 8 days to clinical outcome evaluation at 3, 6, and 12 months, contributing to temporal heterogeneity in reported effects.

Additional heterogeneity arose from differences in patient populations and injury characteristics, including variability in injury level (cervical versus thoracic or lumbar), injury severity (motor-complete versus motor-incomplete SCI), and the inclusion or exclusion of central cord syndrome. Several studies reported subgroup-specific effects, such as greater neurological improvement or biomarker modulation in cervical or motor-complete SCI, while pooled analyses across all SCI subtypes frequently failed to demonstrate statistically significant treatment effects.

Methodological heterogeneity was also evident between clinical and preclinical studies. Animal models employed diverse injury paradigms (compression or hemisection), outcome metrics (behavioral scores, histopathology, biochemical markers), and drug delivery approaches, including nanoparticle-mediated co-delivery systems, which are not directly comparable to human clinical protocols. Collectively, these sources of heterogeneity precluded formal meta-analysis and necessitated a qualitative synthesis of results.

## Discussion

This systematic review and exploratory meta-analysis critically evaluated the available preclinical and clinical evidence regarding the efficacy and safety of minocycline in acute traumatic spinal cord injury (SCI). The principal finding is a clear dissociation between the consistent biological effects observed in experimental models and the limited, heterogeneous, and largely statistically non-significant functional benefits reported in clinical studies. This translational gap has been repeatedly described in SCI research and remains a major obstacle to successful therapeutic development [33].

### Interpretation of Clinical Findings and Quantitative Synthesis

The quantitative synthesis, restricted to two clinical studies with extractable effect estimates (Casha et⍰al., 2012 [21]; Meshkini et⍰al., 2024 [22]), yielded a pooled odds ratio of 1.70 (95% CI 0.95–3.06) for neurological improvement, which did not reach statistical significance. Although the point estimate suggests a possible trend toward benefit, the wide confidence interval reflects limited statistical power and substantial uncertainty. Given the small number of eligible studies, this analysis should be interpreted as exploratory and hypothesis-generating rather than confirmatory.

Several methodological and clinical factors likely contribute to the absence of a statistically robust treatment effect. First, the two studies differed markedly in route of administration and dosing strategy. Casha et⍰al. employed early intravenous minocycline with high loading doses followed by tapering [21], whereas Meshkini et⍰al. used a lower-dose oral regimen over a shorter duration [22]. These pharmacokinetic differences may have resulted in variable central nervous system exposure, thereby attenuating pooled effect estimates. Second, baseline neurological status, injury level, and injury completeness—key determinants of recovery after SCI—were not uniformly stratified or adjusted for across studies, introducing residual confounding that limits comparability.

### Comparison with Previously Published Clinical and Institutional Evidence

The findings of this review are broadly consistent with previously published institutional and trial-based evidence. The phase II randomized controlled trial by Casha et⍰al. demonstrated acceptable safety and modest improvements in ASIA motor scores, particularly in patients with cervical injuries, but these changes did not reach statistical significance [21]. Subsequent prospective cohort analyses derived from the same trial population identified biologically relevant effects, including modulation of cerebrospinal fluid and serum biomarkers associated with axonal injury severity, without consistent translation into durable functional recovery [26,27].

In contrast, the more recent randomized pilot study by Meshkini et⍰al. reported statistically significant improvement in Frankel grade at six months when minocycline was administered in combination with methylprednisolone, compared with methylprednisolone alone [22]. This discrepancy suggests that minocycline may exert greater therapeutic benefit as part of a multimodal neuroprotective strategy rather than as monotherapy. Similar synergistic effects have been reported in experimental nanoparticle-based delivery systems combining minocycline and methylprednisolone, which demonstrated enhanced anti-inflammatory effects and improved histological outcomes in animal models [28,29].

### Quantitative Synthesis of Clinical Outcomes

This meta-analysis synthesizes the only two available human clinical datasets that reported extractable quantitative outcomes on AIS/Frankel grade improvement following intravenous minocycline administration in acute traumatic spinal cord injury (21,22). Both studies demonstrated directionally positive effects favoring minocycline, which is consistent with the biological rationale suggesting that this agent may attenuate secondary injury processes after SCI. Nevertheless, despite encouraging signals from individual trials, the pooled effect estimate did not reach statistical significance (OR = 1.70; 95% CI: 0.95–3.06). The wide confidence interval reflects substantial imprecision arising from the very small number of eligible studies and limited sample sizes. Importantly, the two contributing trials differed in effect magnitude: Casha et al. (21) reported a large but highly imprecise estimate, whereas Meshkini et al. (22) observed a more modest yet statistically significant benefit. When combined, these data yield a positive but non-significant trend, underscoring the instability of pooled estimates derived from minimal evidence bases. Assessment of publication bias was not feasible due to the inclusion of fewer than three studies. Established statistical methods for detecting small-study effects are invalid under such conditions, and although a funnel plot was generated for descriptive purposes, it carries no inferential value. Taken together, these findings suggest a promising yet inconclusive therapeutic signal. While minocycline remains a biologically plausible neuroprotective agent, supported by preclinical research and biomarker modulation observed in human studies (26,27), the current clinical evidence is insufficient to support definitive efficacy claims or routine clinical recommendations. Larger, methodologically rigorous trials are required to determine whether these directionally positive findings translate into clinically meaningful and reproducible neurological benefit.

### Insights from Preclinical Studies and the Translational Gap

Preclinical investigations included in this review consistently demonstrated that minocycline modulates key secondary injury mechanisms following SCI, including neuroinflammation, oxidative stress, and apoptotic signaling pathways [30,32]. However, functional recovery outcomes were variable, particularly in studies evaluating minocycline monotherapy. Notably, a rigorously designed cervical contusion model failed to demonstrate significant neuroprotective or functional benefits despite adequate drug exposure [31]. In contrast, studies combining minocycline with FK506 or methylprednisolone reported more favorable behavioral and biochemical outcomes [30,32].

This pattern mirrors the clinical evidence and underscores a fundamental translational challenge in SCI therapeutics: biological activity does not necessarily equate to meaningful neurological recovery. Differences in injury models, therapeutic windows, outcome sensitivity, and pharmacodynamics between animal experiments and human trials likely contribute to this disconnect. As emphasized in recent narrative reviews, many neuroprotective agents with strong preclinical rationale have failed to demonstrate definitive clinical efficacy in SCI, highlighting the need for more rigorous translational frameworks [33].

### Mechanistic Interpretation

The mechanisms of minocycline’s effects in spinal cord injury are largely derived from preclinical and biomarker-based investigations and provide a biologically plausible, though indirect, framework for interpreting the clinical findings of this review. Experimental studies have consistently demonstrated that minocycline inhibits microglial activation and suppresses NF-κB– mediated inflammatory signaling, resulting in reduced expression of pro-inflammatory cytokines such as TNF-α and IL-6 (15,30,32). These effects appear to mitigate early inflammatory cascades that contribute to secondary tissue damage after SCI.

Downstream consequences of attenuated neuroinflammation observed in animal models include reductions in oxidative stress markers, reflected by improved SOD/MDA ratios, and decreased activation of apoptotic pathways such as caspase-3, ultimately promoting preservation of neural tissue and synaptic architecture (30–32). In preclinical combination studies, particularly those utilizing targeted nanoparticle delivery systems, these molecular and histological benefits were more pronounced, suggesting that drug delivery characteristics may substantially influence mechanistic efficacy (29).

In human studies, mechanistic support remains limited but suggestive. Biomarker analyses demonstrated partial modulation of neuroinflammatory or injury-related markers, including early, non-sustained reductions in serum neurofilament light chain (26) and a significant attenuation of CSF HO-1 elevation at day seven following injury (27). Importantly, these biomarker changes were not consistently accompanied by statistically significant improvements in functional clinical outcomes, underscoring the challenge of translating biological effects into measurable neurological recovery.

Taken together, available evidence indicates that minocycline exerts reproducible anti-inflammatory and neuroprotective effects at the molecular and cellular level, primarily supported by preclinical data. While these mechanisms offer a biologically reasonable explanation for the directionally positive trends observed in some clinical outcomes, they should be interpreted as hypothesis-generating rather than confirmatory. Further studies integrating mechanistic biomarkers with adequately powered functional endpoints are required to clarify whether these biological effects translate into clinically meaningful benefit in patients with acute traumatic SCI.

### Strengths of the Study

This systematic review and meta-analysis were conducted using a rigorous and transparent methodological framework that strengthens the credibility of its findings. A comprehensive literature search was performed across five major electronic databases, PubMed, Scopus, Web of Science, Embase, and the Cochrane Library, in accordance with PRISMA 2020 guidelines (23). The absence of language or publication-date restrictions, together with manual reference screening, maximized coverage and reduced the likelihood of selection bias.

Study screening, full-text selection, data extraction, and risk-of-bias assessment were independently conducted by multiple reviewers, with predefined conflict-resolution procedures. This multi-reviewer approach substantially minimized subjective bias and enhanced methodological reliability. Methodological quality was systematically assessed using Joanna Briggs Institute critical appraisal tools tailored to each study design, providing a structured evaluation of internal validity (Supplementary File 5).

In addition, detailed extraction of population characteristics, injury profiles, intervention protocols, timing parameters, outcome measures, and safety data enabled a nuanced qualitative comparison across heterogeneous studies. Although quantitative synthesis was restricted by the limited number of eligible clinical trials, the application of appropriate random-effects models for pooled analysis and transparent reporting of statistical constraints contributed to an accurate and cautious interpretation of effect estimates.

### Limitations and Implications for Interpretation

Important limitations should also be acknowledged. The clinical evidence base remains limited and heterogeneous, with only two studies eligible for quantitative synthesis, resulting in insufficient power to detect small to moderate treatment effects. Formal assessment of publication bias and between-study heterogeneity was not feasible. Variability in dosing regimens, routes of administration, follow-up duration, and outcome measures further constrained comparability. Additionally, many preclinical studies lacked extractable, drug-specific quantitative outcomes, precluding their inclusion in meta-analysis and limiting cross-species inference.

In much more specific vision: The available evidence is derived from a relatively small number of heterogeneous studies (n⍰=⍰9; total N⍰=⍰512), with only a limited subset focusing exclusively on patients with traumatic spinal cord injury. Considerable variability was observed across studies with respect to methodological design, patient demographics, injury characteristics (including level and completeness), minocycline dosing regimens (100–200⍰mg/day), formulation and route of administration (intravenous versus oral), timing of treatment initiation, duration of therapy, and selection of primary outcomes. These sources of clinical and methodological heterogeneity, together with unmeasured factors such as genetic background or rehabilitation protocols, limit the generalizability of findings and restricted quantitative synthesis to an exploratory level. Follow-up durations were predominantly short to mid-term (up to six months), preventing reliable assessment of long-term efficacy and safety. Moreover, the inclusion of open-label and single-blind designs introduced a moderate risk of performance and detection bias, which should be considered when interpreting reported treatment effects. Only two clinical studies (21,22) provided extractable numerical data suitable for quantitative pooling, resulting in limited statistical power, increased uncertainty in effect estimates, and the inability to formally assess between-study heterogeneity. Standard approaches for evaluating publication bias, including Egger’s regression, Begg’s test, and trim-and-fill, were not applicable due to the small number of studies, and the funnel plot generated was intended for descriptive visualization only. Methodological differences between these two trials, particularly regarding sample size, dosing strategies, and timing of minocycline administration, further contribute to potential inconsistency. Additionally, other clinically relevant neurological outcomes, such as ASIA motor and sensory scores, SCIM, and FIM, could not be pooled because of incompatible reporting formats or insufficient numerical data. Finally, although preclinical investigations of minocycline in SCI are extensive (29–32), most lacked extractable, drug-specific quantitative outcomes, precluding their formal integration into the meta-analysis. Collectively, these minocycline as a routine clinical therapy for spinal cord injury.

These limitations necessitate cautious interpretation of the findings and preclude definitive conclusions regarding clinical efficacy. While minocycline appears safe and biologically active, limitations underscore the substantial gap between current evidence and the establishment of the current evidence does not support its routine clinical use for improving neurological or functional outcomes following acute traumatic SCI.

### Clinical Implications

The available evidence suggests that minocycline is generally well tolerated in patients with acute spinal cord injury and demonstrates biologically neuroprotective effects, primarily supported by preclinical studies and limited human biomarker data (26,27,30–32). However, current clinical evidence remains insufficient to support routine therapeutic use or definitive efficacy conclusions.

Small clinical trials have explored minocycline administration during the acute and subacute phases of SCI, indicating an acceptable safety profile but yielding inconsistent and largely non-significant improvements in functional neurological outcomes, including ASIA motor and sensory scores (21,26). Although experimental data suggest that early modulation of post-injury neuroinflammation may be beneficial, the optimal therapeutic window, dosing strategy, and duration of treatment have not been established in humans.

Given the absence of clarifying, confirmatory randomized evidence, minocycline should presently be regarded as an investigational or adjunctive therapeutic candidate rather than a standard component of SCI management. Its use outside clinical trial settings should be guided by institution-specific protocols and careful risk–benefit assessment. The integration of minocycline with existing clinical strategies, including surgical decompression, hemodynamic optimization, and rehabilitation, remains exploratory and requires validation in adequately powered, controlled studies.

Overall, while minocycline represents a promising candidate targeting secondary inflammatory injury mechanisms in SCI, substantial gaps persist between current exploratory evidence and widespread clinical application. Larger, methodologically rigorous randomized trials are necessary before minocycline can be recommended as part of routine clinical practice.

### Future Directions

Targeted and well-designed research is essential to translate the potential of minocycline into a definitive treatment for acute spinal cord injury (SCI). To establish efficacy and define optimal dosing and therapeutic windows, large-scale, multicenter, double-blind Phase III randomized controlled trials (RCTs) in homogeneous SCI populations are required. Evaluating combination therapies, such as minocycline with methylprednisolone, other neuroprotective agents, or intensive neurorehabilitation, represents a logical next step. Studies should also assess long-term outcomes (1–2 years) including functional status, quality of life, and late adverse effects. Cost-effectiveness analyses and investigations into predictive biomarkers (e.g., serum NfL, cytokine profiles) are critical for identifying responsive patient subgroups and optimizing treatment strategies.

Future research should prioritize adequately powered, multicenter RCTs with standardized intervention protocols to enable valid meta-analytic synthesis. Harmonized reporting of AIS grade, ASIA motor and sensory scores, and functional recovery measures such as SCIM and FIM is essential. Long-term follow-up and stratification by injury level and severity will further strengthen clinical conclusions.

Preclinical studies should provide transparent, quantitative reporting of minocycline-specific outcomes to facilitate translational analyses. Investigations into optimal dosing, therapeutic time windows, and interactions with surgical or rehabilitation strategies are also warranted.

Given the promising yet statistically inconclusive findings, further rigorous clinical trials are needed to clarify the therapeutic potential of minocycline in acute SCI.

## 5. Strengths of the Study

This systematic review and meta-analysis were conducted using a rigorous and transparent methodological framework that strengthens the credibility of its findings. A comprehensive literature search was performed across five major electronic databases, PubMed, Scopus, Web of Science, Embase, and the Cochrane Library, in accordance with PRISMA 2020 guidelines (23).

The absence of language or publication-date restrictions, together with manual reference screening, maximized coverage and reduced the likelihood of selection bias.

Study screening, full-text selection, data extraction, and risk-of-bias assessment were independently conducted by multiple reviewers, with predefined conflict-resolution procedures. This multi-reviewer approach substantially minimized subjective bias and enhanced methodological reliability. Methodological quality was systematically assessed using Joanna Briggs Institute critical appraisal tools tailored to each study design, providing a structured evaluation of internal validity (Supplementary File 5).

In addition, detailed extraction of population characteristics, injury profiles, intervention protocols, timing parameters, outcome measures, and safety data enabled a nuanced qualitative comparison across heterogeneous studies. Although quantitative synthesis was restricted by the limited number of eligible clinical trials, the application of appropriate random-effects models for pooled analysis and transparent reporting of statistical constraints contributed to an accurate and cautious interpretation of effect estimates.

## Conclusion

This systematic review and exploratory meta-analysis highlight a clear dissociation between the robust biological effects of minocycline observed in preclinical models and the limited and inconsistent clinical benefits demonstrated in human studies of acute traumatic spinal cord injury. While minocycline appears to be safe and biologically active, particularly in modulating inflammatory and oxidative pathways, current clinical evidence does not support its routine use for improving neurological or functional outcomes. The findings underscore the need for adequately powered, methodologically rigorous randomized trials with standardized outcome reporting and long-term follow-up to determine whether minocycline can meaningfully contribute to therapeutic strategies for acute SCI.

## Data Availability

All data produced in the present study are available upon reasonable request to the authors

## Abbreviations

AIS: ASIA Impairment Scale
ASIA: American Spinal Injury Association
BBB: Basso, Beattie, and Bresnahan locomotor rating scale
CI: Confidence Interval
CSF: Cerebrospinal Fluid
CXCL10: C-X-C motif chemokine ligand 10
FIM: Functional Independence Measure
FND: Functional Neurologic Decline
FK506: Tacrolimus
GSH: Glutathione
HO-1: Heme Oxygenase-1
IL-1β: Interleukin-1 beta
IL-6: Interleukin-6
IP: Intraperitoneal
IV: Intravenous
JBI: Joanna Briggs Institute
MC: Minocycline
MDA: Malondialdehyde
MMP-9: Matrix Metalloproteinase-9
MPO: Myeloperoxidase
MP: Methylprednisolone
NfH: Neurofilament Heavy Chain
NfL: Neurofilament Light Chain
NCAM: Neural Cell Adhesion Molecule
NOx: Nitric Oxide Metabolites
OR: Odds Ratio
PO: Per os (oral administration)
RCT: Randomized Controlled Trial
SCI: Spinal Cord Injury
SCIM: Spinal Cord Independence Measure
SOD: Superoxide Dismutase
TBI: Traumatic Brain Injury
TNF-α: Tumor Necrosis Factor alpha

## Author Contributions

FF and FT developed the inclusion/exclusion and data extraction sheets, conducted the analysis, and oversaw the review process. AG, MM, and MF conducted title and abstract screening. RQ and ME performed full-text screening. MH and AS conducted data extraction. MJ and MM assessed methodological quality and risk of bias. MJ assisted with risk of bias assessment. MG, HM, SBK, and SO verified data extraction and organized supplementary files. SY and AZ edited the manuscript and provided critical revisions.

## Declaration of AI Use

Artificial intelligence (AI) tools (ChatGPT) were used to improve the grammar, clarity, and language of this manuscript. The authors reviewed and approved all content, and take full responsibility for the integrity and accuracy of the work.

## Acknowledgments

The authors have no acknowledgments to declare.

## Appendix and Supplementary Material

- Supplementary File 1. PRISMA checklist
- Supplementary File 2. Systematic Review Protocol
- Supplementary File 3. Search Strategy
- Supplementary File 4. Excluded studies
- Supplementary File 5. Full Text screening
- Supplementary File 6. Data extraction Sheets
- Supplementary File 7. Risk of bias assessment

